# Epidemiological model for the inhomogeneous spatial spreading of COVID-19 and other diseases

**DOI:** 10.1101/2020.07.08.20148767

**Authors:** Yoav Tsori, Rony Granek

## Abstract

We suggest a mathematical model for the spread of an infectious disease in human population, with particular attention to the COVID-19. Common epidemiological models, e.g., the well-known susceptible-exposed-infectious-recovered (SEIR) model, implicitly assume fast mixing of the population relative to the local infection rate, similar to the regime applicable to many chemical reactions. However, in human populations, especially under different levels of quarantine conditions, this assumption is likely to fail. We develop a continuous spatial model that includes five different populations, in which the infectious population is split into latent (or pre-symptomatic) and symptomatic. Based on nearest-neighbor infection kinetics, we arrive into a “reaction-diffusion” model. Our model accounts for front propagation of the infectious population domains under partial quarantine conditions, which is present on top of the common local infection process. Importantly, we also account for the variable geographic density of the population, that can strongly enhance or suppress infection spreading. Our results demonstrate how infected domains spread outward from epicenters/hotspots, leading to different regimes of sub-exponential (quasi linear or power-law) growth. Moreover, we show how weakly infected regions surrounding a densely populated area can cause rapid migration of the infection towards the center of the populated area. Predicted heat-maps show remarkable similarity to recently media released heat-maps. We further demonstrate how localized strong quarantine conditions can prevent the spreading of the disease from an epicenter/hotspot, significantly reducing the number of infected people. Application of our model in different countries, using actual demographic data and infectious disease parameters, can provide a useful predictive tool for the authorities, in particular, for planning strong lockdown measures in localized areas.

## I. INTRODUCTION

The COVID-19 pandemic is now spread over most of the globe. Its vast consequences are associated with severe public health issues, i.e. overwhelmed health system and high death toll, and a huge economic crisis worldwide [1–5]. In order to optimize decisions in both aspects, health and economy, governments need information and predictions about the spatial distribution of the disease [6], thereby allowing selective quarantine or lockdown measures [7, 8].

Infectious disease spreading models are largely based on the assumption of perfect and continuous “stirring”, similar to the one used to describe the kinetics of spatially-uniform chemical reactions. In particular, the well-known susceptible-exposed-infectious-recovered (SEIR) model, builds on this assumption. Early in the COVID-19 pandemic spread, different modeling groups have used such models to predict the epidemic evolution in China and in other countries [9–11]; these predictions urged WHO to issue a global warning. Wu *et al*. were the first to model the COVID-19 spreading [9]. They applied the SEIR model based on data from the very early (exponential) stage of the outbreak, to predict epidemic spread mainly in Wuhan and mainland China. Extensions for this first attempt were quick to follow. Ivorra and Ramos applied the “Be-CoDiS” mathematical model – a multi-population extension of the SEIR model – to COVID-19 [12, 13]. Fitting the parameters of the latter model to a longer period of evolution, up to the time the outbreak nearly peaked (maximum number of new daily infected people), yielded remarkably accurate predictions for the following stages. More recently, He *et al*. [14] and Giordano *et al*. [15] provided further improvements and analysis on the original application of the SEIR model to COVID-19 [9].

As mentioned, conventional epidemiological models assume spatially uniform (statistical) frequency of encounters between infectious and susceptible people, which is associated with uniform (spatial) densities of these populations at all times [9, 16]. As such, these models do not require any spatial variable. However, the assumption of “infinitely fast mixing” might fail even in normal life conditions, let alone under (the often used) various gathering restrictions or moderate quarantine conditions [7]. As a consequence, the predictions of such models are relatively poor, and often require refitting the infection rate constant as the epidemic progresses. Specifically, these models usually fail to predict correctly a cross-over from the initial exponential growth of the number of infected people to sub-exponential growths which might even be a power-law behavior [17–19]. Early data from China and Italy demonstrated such a wide temporal regime of quasi power-law growth, which occurred much prior to the peak of the epidemic [20–22]. We hypothesize that such a temporal behavior results partially from “front propagation” of infected geographical domains, similar to the spreading of wildfire.

In this paper, we develop a novel model for viral infection. Our model improves on the SEIR model, in particular in the context of the COVID-19 pandemic, in four major aspects. The first is the derivation of a spatial spreading, diffusion-like, operator that takes account of the front propagation of the epidemic. The second, and strongly linked to the first, is the ability to account for the geographical population density variation and study its effect on the spreading. The combination of these two aspects leads to spreading of the disease into densely populated areas. The third aspect is the account of geographic variation in quarantine levels if such are employed. Lastly, for COVID-19 we split the infectious population into a transient “infectious-asymptomatic” group and an “infectious-symptomatic” group. Our results indeed demonstrate unique features of the disease spreading depending on the spatial dependence of population density, location of the initial “hotspots”, and spatial variation of the quarantine levels. They show that the accumulated number of infected people manifests major qualitative and quantitative deviations from the classical (yet generalized to five populations) SEIR model.

### A. Generalized SEIR model

Our model, which stems from the SEIR model, includes five populations associated with different stages of the disease. Furthermore, we account for the spatially varying density of people, *n*(**x**), between different areas of the geographical region under study, which assumed in the present study disconnected from other regions. Our model also aims to predict the effect of different quarantine levels imposed in different areas within the region of study, which is modeled via *spatial dependence* of infection rates.

Let *h* (**x**, *t*) be the 2D (areal) concentration of susceptible (healthy but not immune) people, *b* (**x**, *t*) – the concentration of infected people that do *not* yet infect others and are *not* yet symptomatic (i.e within the incubation period [23]), *w* (**x**, *t*) – the concentration of infected people that can already infect others but are still asymptomatic (i.e. still within the incubation period), *f* (**x**, *t*) – the concentration of infected and symptomatic people, and *r* (**x**, *t*) – the concentration of people that have recovered from illness, thus assumed (here) to be immune from a second infection. We require that the total *local* concentration of people is equal to prescribed values *n*(**x**) at different positions **x** (obtained, e.g., from public databases) and *independent of time*:

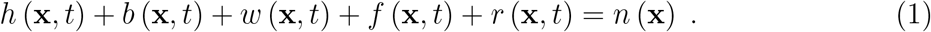

It follows that the total population number, *N* = ∫_area_ *d*^2^*x n* (**x**), is also not changing with time. We note in passing that, if spatial dependence could be ignored, these variables correspond to the variables of the well-known SEIR epidemiological model [16, 24, 25] as follows: *h* ↔ *S* (susceptible), *b* ↔ *E* (latent), *w* + *f* ↔ *I* (infectious), *r* ↔ *R* (removed). Unlike in the SEIR model, in our model the infectious population (*I*) is split into two populations, symptomatic (“sick”, *f*) and pre-symptomatic (*w*). In what follows, capital letters will denote the corresponding global quantities, e.g., *H*(*t*) = ∫_area_ *d*^2^*x h* (**x**, *t*), *B*(*t*) =∫ _area_ *d*^2^*x b* (**x**, *t*), etc.

In order to develop the (continuous) space epidemic spread model, we consider first a (2D) discrete space, in which the nodes are defined as either homes, building apartments, or units of area (e.g., quarantine units of 10^4^m^2^), and so on. We define by *h*_*i*_(*t*) the number of susceptible people at node *i* at time *t*. Similarly *b*_*i*_(*t*), *w*_*i*_(*t*), *f*_*i*_(*t*), and *r*_*i*_(*t*) describe the numbers of the different populations at each node. Infection can occur with a rate constant *k*_1_(*i*) when an infectious person and a susceptible person meet at the same node *i*, and at a rate constant *k*_2_(*j, i*) = *k*_2_(*i, j*) when infectious and susceptible persons meet at nearest-neighbor (NN) nodes *i* and *j* (independent whether the infectious person is at *i* and susceptible is at *j* or *vice versa*). The total number of people at node *i* is denoted by *n*_*i*_. Accordingly, the set of master equations for the distribution of these populations is

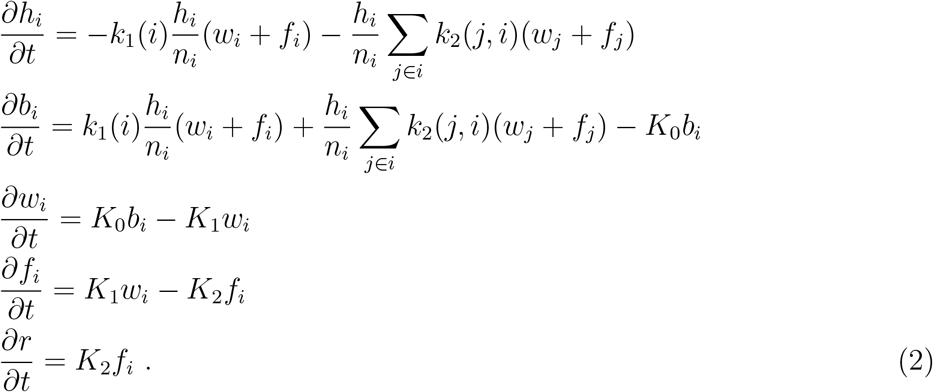

where *j ∈ i* stands for node *j* that is NN to *i*. In Eq. (2), *K*_0_, *K*_1_, and *K*_2_ are the rate coefficients for the transformation of the *b*-population to *w*, of *w* to *f*, and of *f* to *r*, respectively.

We now transform the master Eq. (2) to the continuum using the Kramers-Moyal expansion [26], **x** ↔ *i*, in which each node is assumed to occupy and area *δ*^2^ where *δ* is the grid spacing (lattice constant). Using the symmetry for infection rates between NN nodes *i* and *j, k*_2_(*j, i*) = *k*_2_(*i, j*), and defining the local *concentration* (i.e. number density) of a population *y* as *y* (**x**, *t*) *≡ y*_*i*_(*t*)*/δ*^2^, we obtain

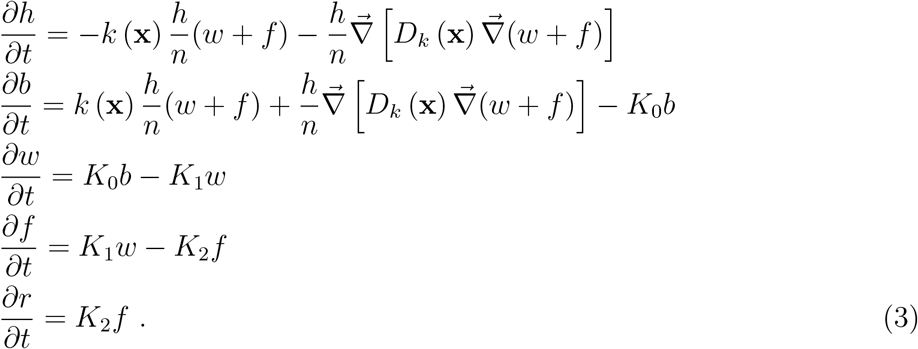

where the last line in Eq. (3) can be replaced – using the conservation law Eq. (1) – by *r* = *n* − (*h* + *b* + *w* + *f*). In Eq. (3), *k* (**x**) = *k*_1_ (**x**) + *zk*_2_ (**x**) defines an effective local rate coefficient of infection growth, and *D*_*k*_ (**x**) = *k*_2_ (**x**) *δ*^2^ is an effective diffusion coefficient of the infection spreading; *δ* is the inter-node spacing, and *z* is the coordination number of the grid. The gradient term in the first two equations describes “front propagation” and domain growth of infectious people under quarantine conditions. Note that for the case of homogeneous distribution of all populations and homogeneous rate constants, *k* can be identified as the SEIR parameter ratio *R*_0_*/τ*_*I*_, where *R*_0_ is the basic reproductive number and *τ*_*I*_ is the mean infectious period. The latter can be related to our rate coefficients *K*_1_ and *K*_2_ by 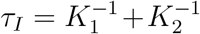. It is easy to verify by summing all lines in Eq. (3) that *∂n* (**x**, *t*) */∂t* = 0, as required. Thus, any initial inhomogeneous population density distribution *n* (**x**) is not altered by our infection spreading model.

For simulation purposes we rescale the local densities by the mean total population density (in the whole region under study), *n*_0_, such that 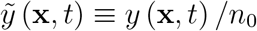. In particular, *ñ* (**x**) = *n* (**x**) */n*_0_ presents the relative local population density. In addition, distance is scaled by *δ*, i.e. 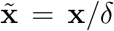, such that 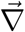 becomes dimensionless. This leads to the following scaled equations

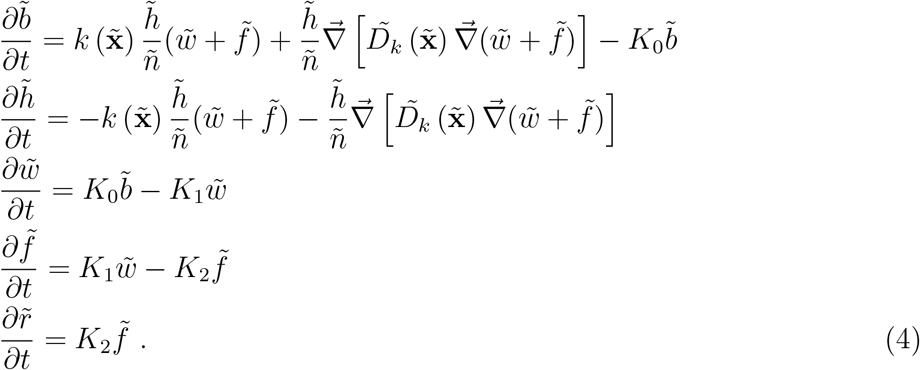

where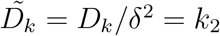

The parameters to be used for COVID-19 pandemic should be obtained from the up-to-date literature. In the absence of any quarantine conditions or safety measures (e.g., use of masks), *R*_0_ = 2.5 and *τ*_*I*_ = 16.6 days [27] and thus the growth rate *k* = *R*_0_*/τ*_*I*_ is about 0.15 days^−1^. The rate *k* can be reduced only by reduction of *R*_0_, i.e. putting in place quarantine/safety measures. The mean time for symptoms to appear (from the moment of infection), *τ*_*S*_, is known to be about 5 days (ranging between 2-11 days) which sets up 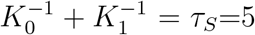 days. However, *K*_0_ itself, describing the transition from “infected but non-infecting” to “asymptomatic-infecting”, is not reported; a reasonable guess would be *K*_0_ ≃ 1*/*(2 days) [28]. This is supported by the hypothesis that people are infecting already about 2-3 days before they show symptoms; therefore a reasonable choice would be 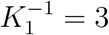 days, in agreement with 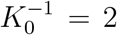 days. The rate coefficients *K*_1_ and *K*_2_, describing the transitions from asymptomatic-infecting to symptomatic-infecting, and from symptomatic-infecting to recovered, respectively, must obey 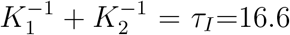 days (i.e. the whole infection period), implying 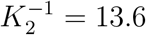 days. The dimensionless effective diffusion coefficient 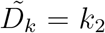 is the most difficult model parameter to estimate and is sensitive to the choice of nodes. Likely 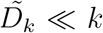, since *k* = *k*_1_ + *zk*_2_ and we may also assume *k*_2_ ≤ *k*_1_ and *z* ≃ 4, 6 (square and hexagonal lattices). For numerical purposes in the present study study we chose *k*_1_ = *k*_2_ = 0.03, and *z* = 4 yielding *k* = 0.15 and 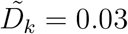.

In the proceeding section we solve this spatially dependent multiple population model, at different initial conditions and parameters, and use a few specific inhomogeneous density populations *n* (**x**) as an example. Obviously, for realistic predictions one requires: (i) detailed local population density data (i.e. density maps), and (ii) data for the initial local densities of the above five different populations (i.e. “heat-maps”), both given to the grid size resolution – requiring cooperation with authorities. The present work is therefore limited to present the strength of the model and its ability to give insight on the way the infection spreads under different levels and spatial variation of quarantine. For brevity, henceforth, we drop the ‘∼’ sign from the notations of (density) normalized spatially dependent variables, i.e.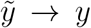. Recall that capital letters denote *global* quantities, i.e. spatial integrals of the lowercase spatially dependent quantities, e.g., *F* = ∫ *f*(**x**)*d*^2^*x*, representing now the *fraction* of the specific population – out of the total population – as now *f* (**x**) implies 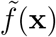.

## II. RESULTS

The initial conditions of an epidemic are unknown unless in exceptional cases, yet they have major consequences on the number of infected people. In all examples below, we shall use identical initial conditions for the global quantities as follows: *W* = *R* = *F* = 0 and *B* = 10^−3^. In our non-uniform model, we are able to analyze the effect of the different, non-uniform, initial conditions, yet identical global initial conditions. Comparing the evolution in time of the global quantities, between the locally different – yet globally identical – initial conditions, we will assess both the spreading patterns and the overall effect of the epidemic.

Before we investigate the importance of population density heterogeneity, we analyze the model results in the special case where all population densities are kept uniform at all times. In this limit, our model converges to an effective SEIR-type model, with just a single additional population. It should be emphasized that the SEIR-type limit cannot be achieved realistically as it requires rapid mixing of people, and is studied here only for comparison. Obviously, in this case of uniform distributions, the global (spatial integral) quantities do not provide additional information.

Figure 1 shows the model predictions when *n, b, h, w, f*, and *r* are spatially uniform and for the above stated initial conditions (*W* = *R* = *F* = 0 and *B* = 10^−3^). Figure 1(a) shows the time evolution of the model variables *H, B, W, F*, and *R*. At a time *t* ≃ 90 days the epidemic attains its peak, i.e. the number of symptomatic people (*F*) attains a maximum (blue curve) and later declines. The number of recovered/removed people, *R* (green curve), is increasing. In this particular example, at very long times *R* attains a value of *R* ≈ 90% and correspondingly *H* ≈ 10%. Thus, within our five-population SEIR-type model, and the parameters chosen, “herd immunity” is reached at 90% infection.

**FIG. 1.**
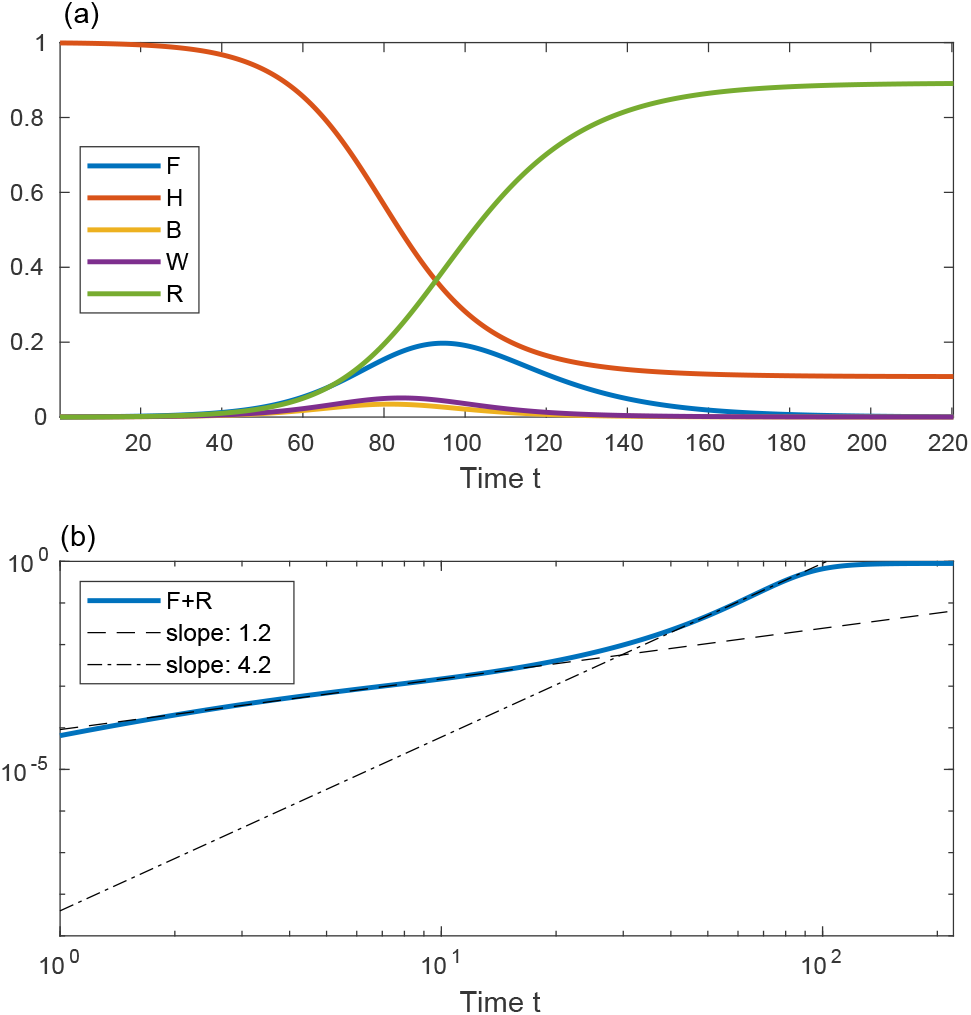
(a) Solution of the epidemic model for the case of spatially uniform population densities *n, b, h, w, f*, and *r*. All curves depict the global population quantities (capital letters), amounting here to simple multiplication of the local densities by the area. The initial conditions are *B* = 10^−3^ and *W* = *F* = *R* = 0. (b) A log-log plot of the total symptomatic and recovered populations (*F* + *R*) *vs* time (in days). The dashed and dash-dotted lines are fits at *t* = 10 and *t* = 73, respectively. In this and in all other figures *D*_*k*_ = 0.03, *k* = 0.15 days^−1^, *K*_0_ = 1*/*2 days^−1^, *K*_1_ = 1*/*3 days^−1^, *K*_2_ = 1*/*13.6 days^−1^.

A common quantity used to follow the epidemic is the number of people that have been infected until time *t*. If we limit the analysis to only those that showed symptoms, this number is the sum of the total symptomatic and recovered populations, *F* + *R*. In Fig. 1 (b) we show *F* + *R vs* time on a log-log scale. We observe a separation of the growth into two power-law regimes, with the early evolution exponent ≃ 1.2 being much smaller than the late evolution exponent ≃ 4.2.

Let us now look at epidemic evolution that initiates from different nonuniform initial conditions, in either uniformly or non-uniformly populated area. Consider, first, several infection centers of the latent *b*-population, randomly distributed within a uniformly populated area, Fig. 2 (a), *t* = 0. Although *n* is uniform at all times, all specific populations are nonuniform at *t >* 0 (even when *h, w, f*, and *r* are set uniformly to zero at *t* = 0). The integrated value of *b* is therefore the same as in Fig. 1, i.e. *B* = 10^−3^, and obviously all other populations vanish at *t* = 0. Panels (b)-(f) show the time evolution of the symptomatic population *f*. As can be seen, each infection point grows locally and develops into a “ring” centered around the original point. These rings further grow and coalesce into a complex boundary pattern which keeps evolving. Figure 3 shows the integrated values of *F, H, B, W*, and *R* and *F* + *R*. Compared to Fig. 1 for the case of uniform *b*, here the epidemic peak (maximum in the *F* -population) occurs at a longer time and, more importantly, is significantly smaller – an effect often termed as “curve flattening”. The early evolution power-law exponent is similar to the uniform *b* case, however, the long-time exponent ≃ 2.8 is significantly smaller, leading to a significantly longer saturation time where”herd-immunity” is reached.

**FIG. 2.**
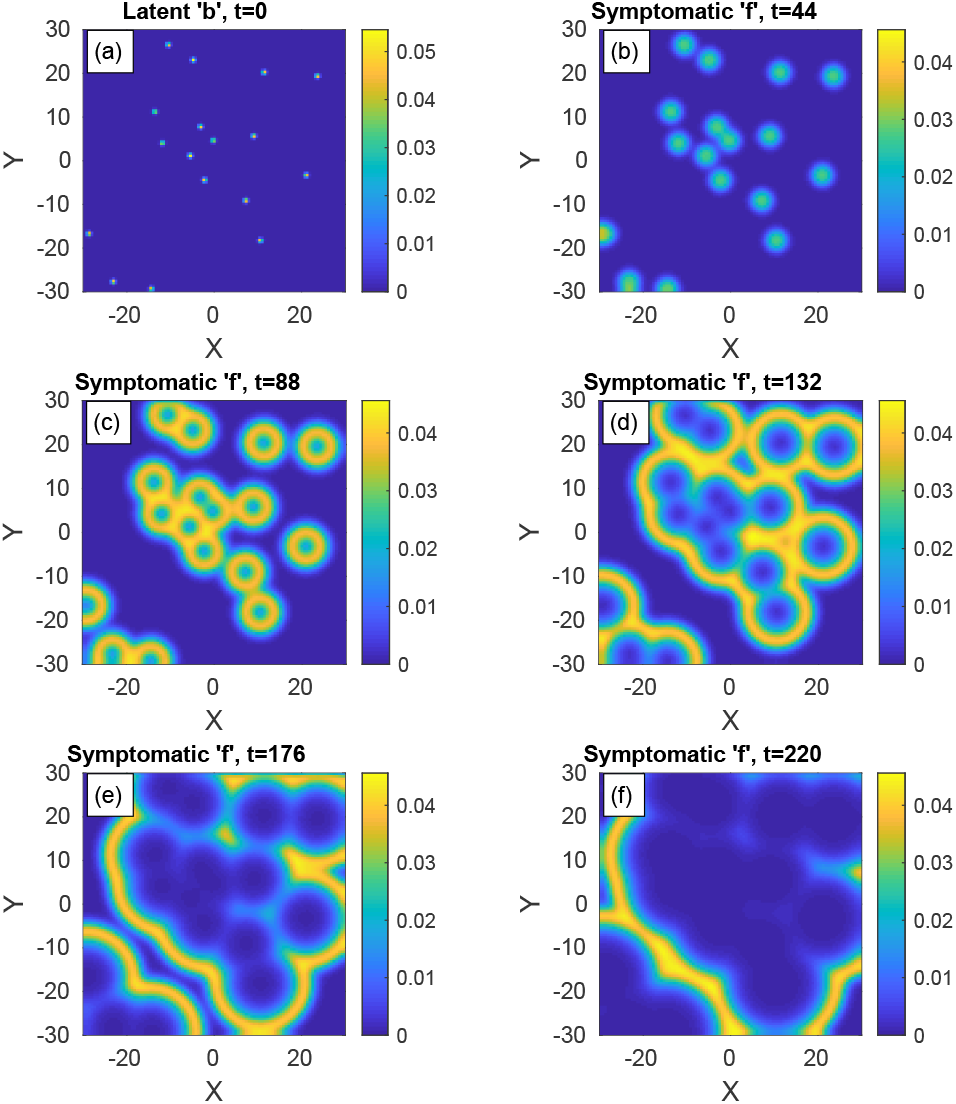
Time evolution of an epidemic with spatially nonuniform infection centers. (a) At *t* = 0 (top-left panel), there are small infection centers of the latent population ‘*b*’ distributed randomly in space. The integrated value of *b* is the same as in Fig. 1, i.e. *B* = 10^−3^. *n* is uniform and all other populations are set initially to zero: *w* = *f* = *r* = 0. Panels (b)-(f) show the spread of the symptomatic population ‘*f* ‘as time progresses.

**FIG. 3.**
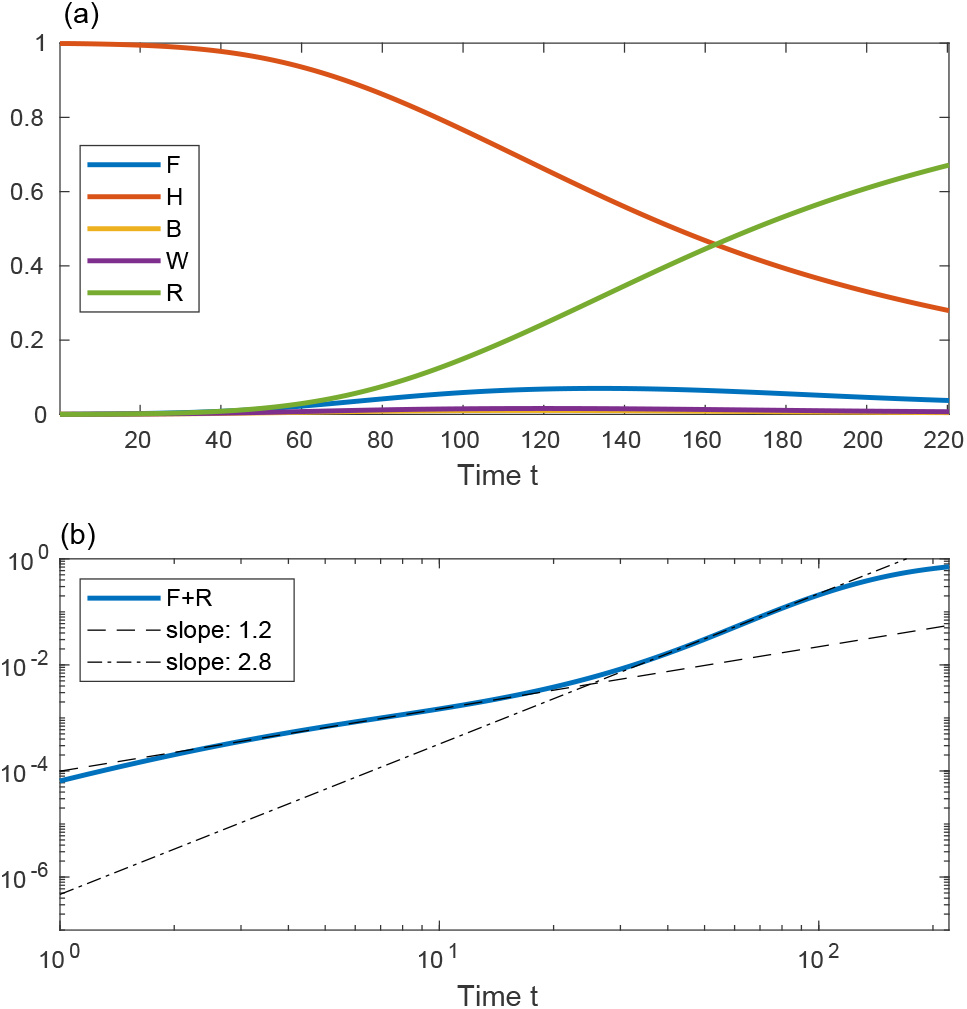
(a) Global population curves *F, H, B, W*, and *R vs* time in days corresponding to the random infection centers in Fig. 2. (b) *F* + *R vs* time in with log-log axes. Dashed and dash-dot curves are linear fits at *t* = 10 and *t* = 73, respectively.

Consider a rather different initial spreading of the *b*-population, still with *n* uniform in space. In Fig. 4 the initial conditions are two relatively large infection centers, instead of many small ones as in Fig. 2. These centers grow and develop into ring-like structures, later start to overlap, and subsequently merge into one big ring that continuously spreads outwards. The core of the rings is seen to evolve quickly to contain mostly recovered population (since *r* ≃ *n* − *f*).

**FIG. 4.**
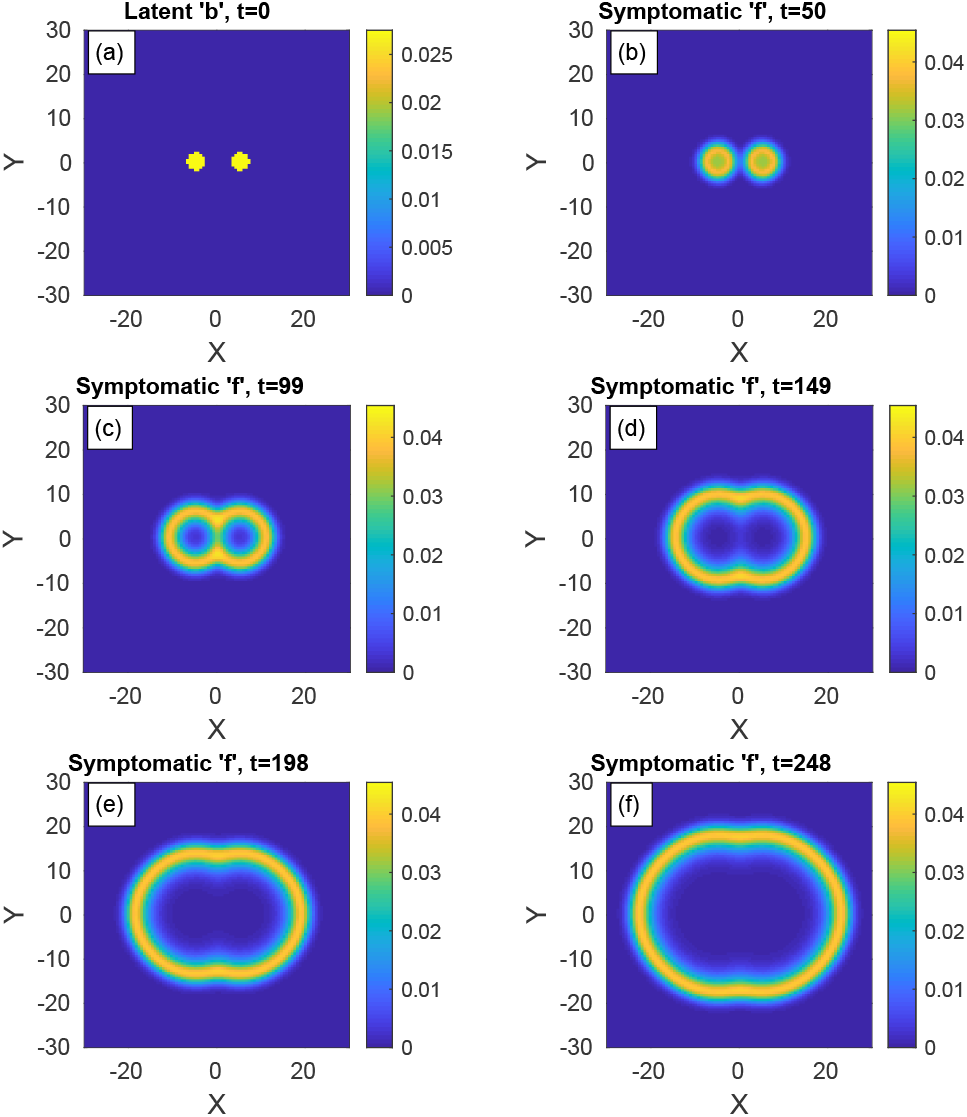
Time evolution of an epidemic starting from two infection centers. (a) Initial conditions of *b. B* – the global value of *b* – is the same as in Figs. 1-3. *n* is uniform and all other populations are initially zero: *w* = *f* = *r* = 0. Panels (b)-(f) depict the spreading pattern of the symptomatic population ‘*f* ‘as time progresses. The two circular domains grow and merge into one oval-like domain.

In Fig. 5 we show the evolution in time of global populations corresponding to the evolution patterns depicted in Fig. 4. Comparing to the above cases of uniform (Fig. 1) or random (Figs. 2-3) initial infection centers *b*, here the peak of the epidemic occurs at much longer times and is much smaller in magnitude. The early-time power-law regime is much shorter and quickly crosses over (at *t* = 10) to the long-time power-law behavior with exponent ≃ 2. The latter exponent signifies the 2D front propagation of the domains, whose core corresponds to mostly recovered population (*r*) and ring corresponding to mostly symptomatic population (*f*). It implies that the front is moving with constant velocity, leading to the ∼ *t*^2^ domain growth.

**FIG. 5.**
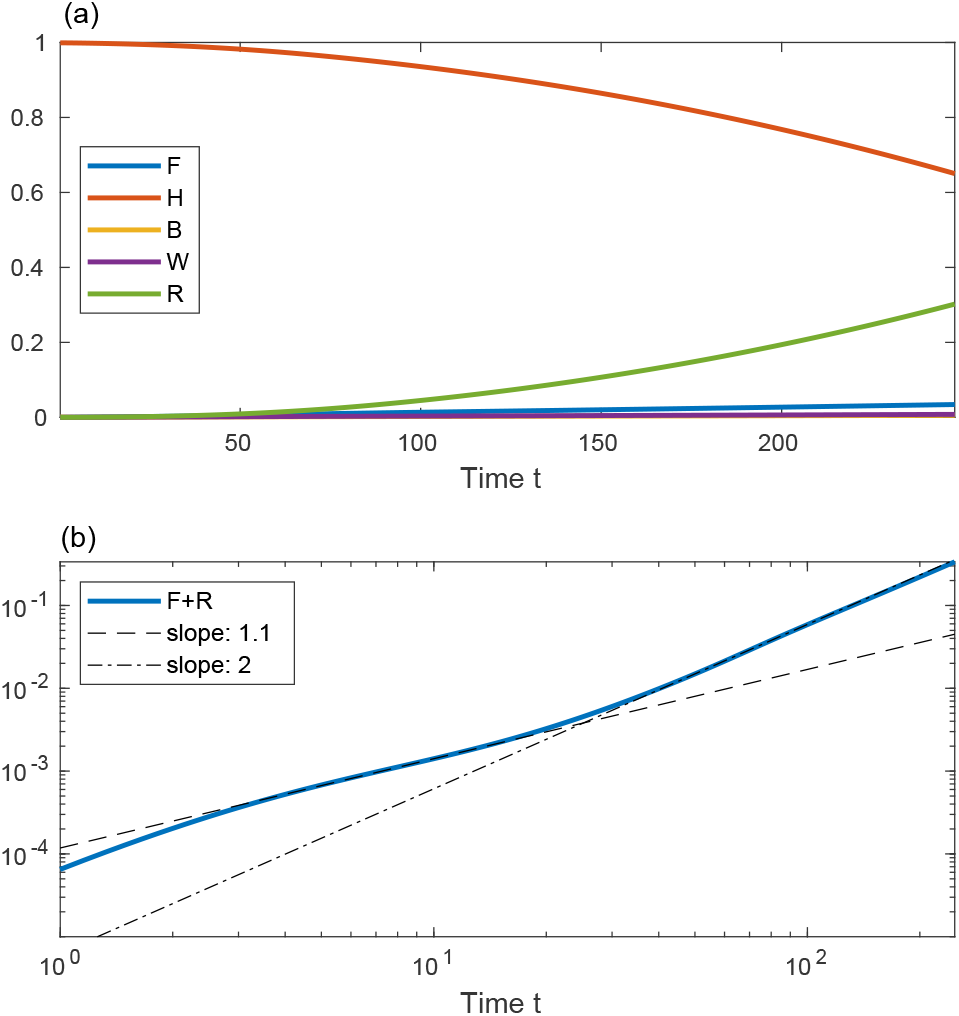
Global population curves *vs* time (in days) corresponding to the circular infection centers in Fig. 4. Compare to the cases of uniform (Fig. 1) or random (Figs. 2-3) distributions of ‘*b*’. Dashed and dash-dotted curves are linear fits at *t* = 10 and *t* = 83, respectively.

We now turn to study situations in which the given population density is not uniform in space, and our model is highly suitable to handle such cases. As case studies, we mimic large density variations, such as those created by cities surrounded by suburban areas. In Figs. 6, 7 and 8 we examine the outbreak near a highly populated circular “city” whose population declines from its center as a Gaussian whose standard deviation (width) is denoted by *ℓ*. We consider here three different cases for the initial conditions: (i) the infection (*b*-population) initiates from inside the city (Fig. 6), (ii) the infection evolves from several, randomly distributed, centers in the city periphery (Fig. 7), and (iii) the infection evolves from two major centers in the city periphery (Fig. 8). In all figures, the population heat-maps are on the left-hand-side, and the corresponding global quantities are depicted on the right-handside.

**FIG. 6.**
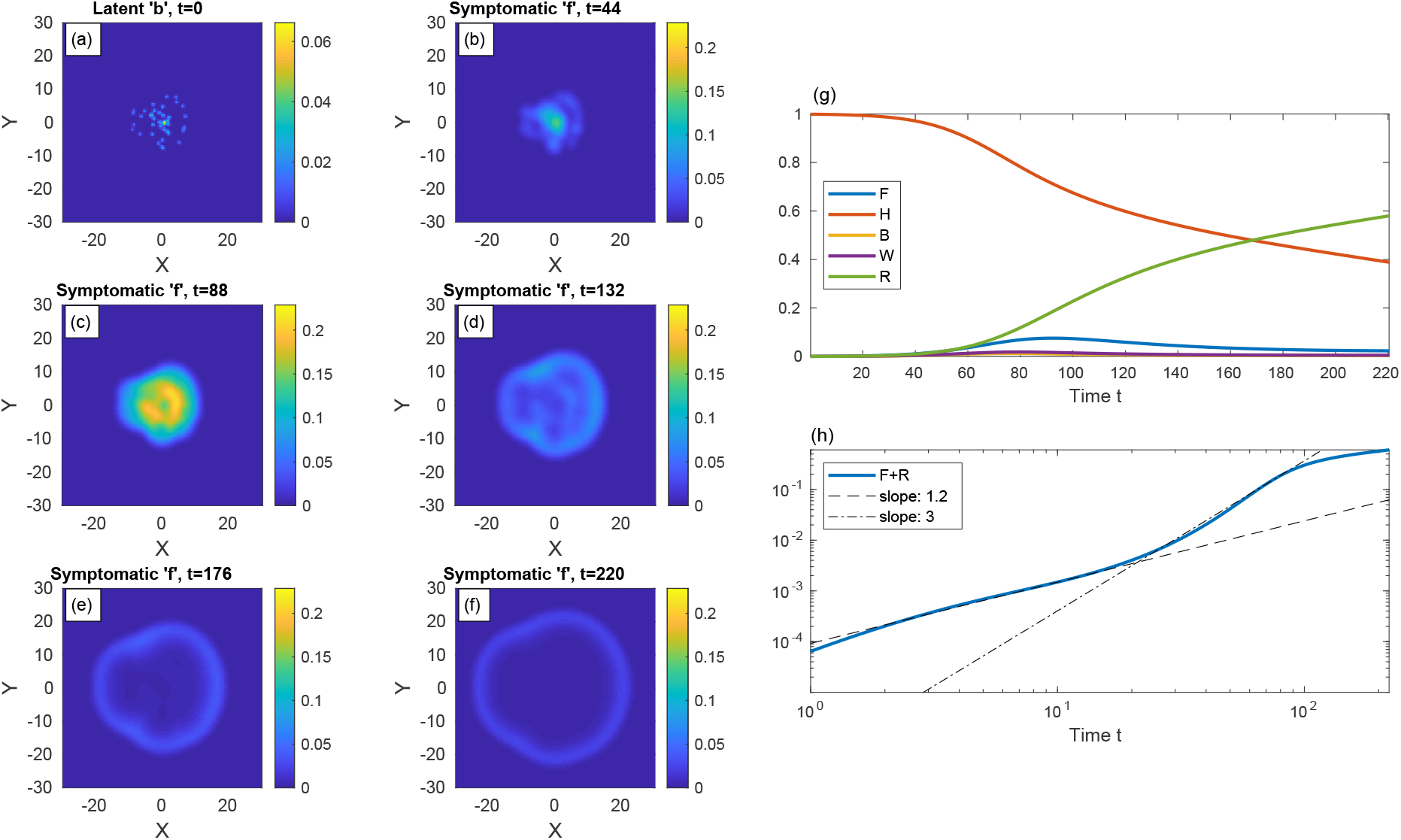
Time evolution of an epidemic starting from multiple infection centers inside a heavily populated region (“city”, panel (a)). The population density of the city *n* is nonuniform and given by 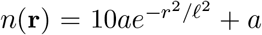, with **r** = (*x, y*), *ℓ* = 10, and *a* taken such that the spatial average of *n* is 0.2. The integrated value of *b* is the same as in Figs. 1-3. All other populations are initially zero: *w* = *f* = *r* = 0. Panels (b)-(f) show the spread of the symptomatic population ‘*f* ‘as time progresses. The global populations and the sum *F* + *R* are shown in (g) and (h), respectively.

**FIG. 7.**
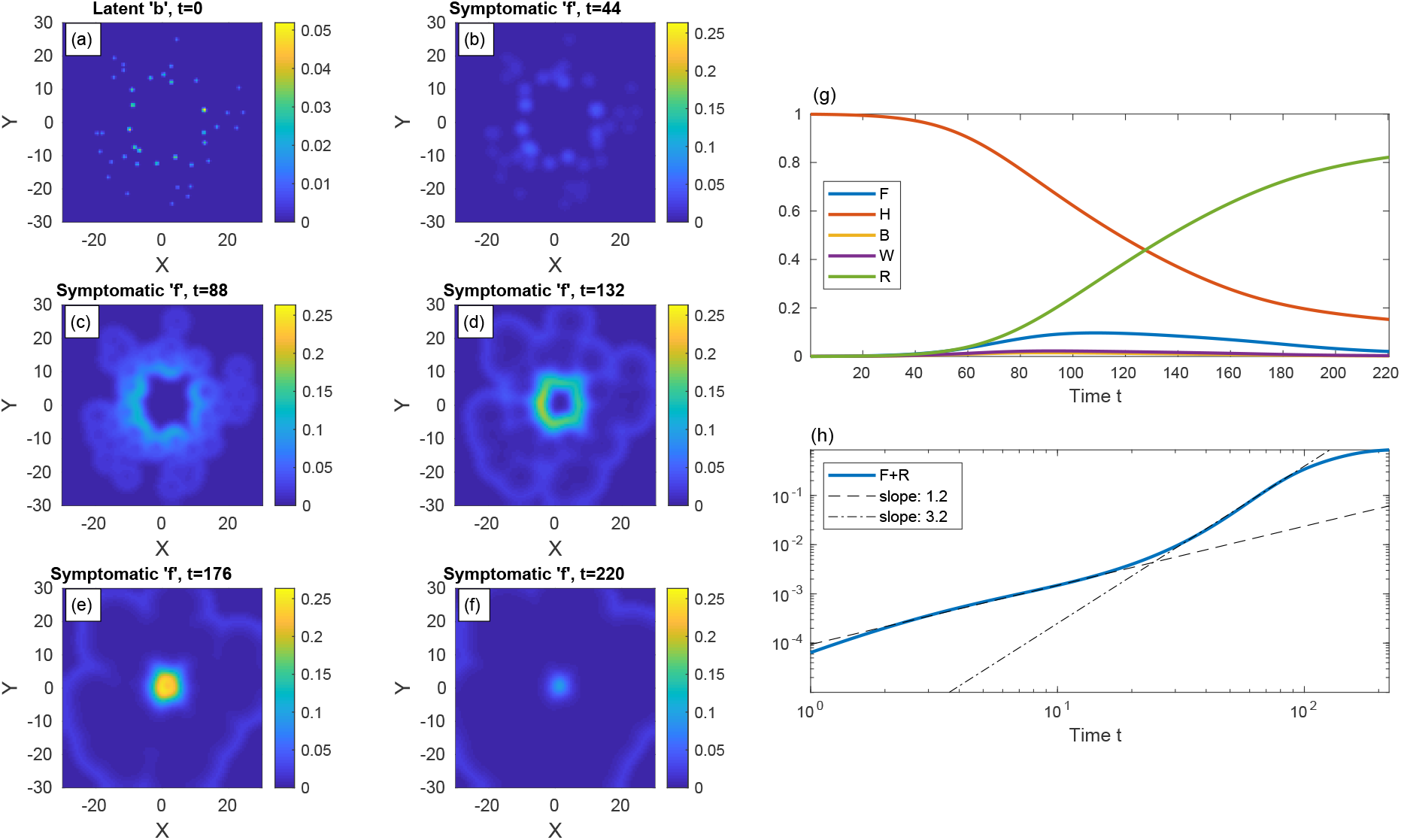
The same as in Fig. 6 but with the infection centers at *t* = 0 located outside of the “city”.

**FIG. 8.**
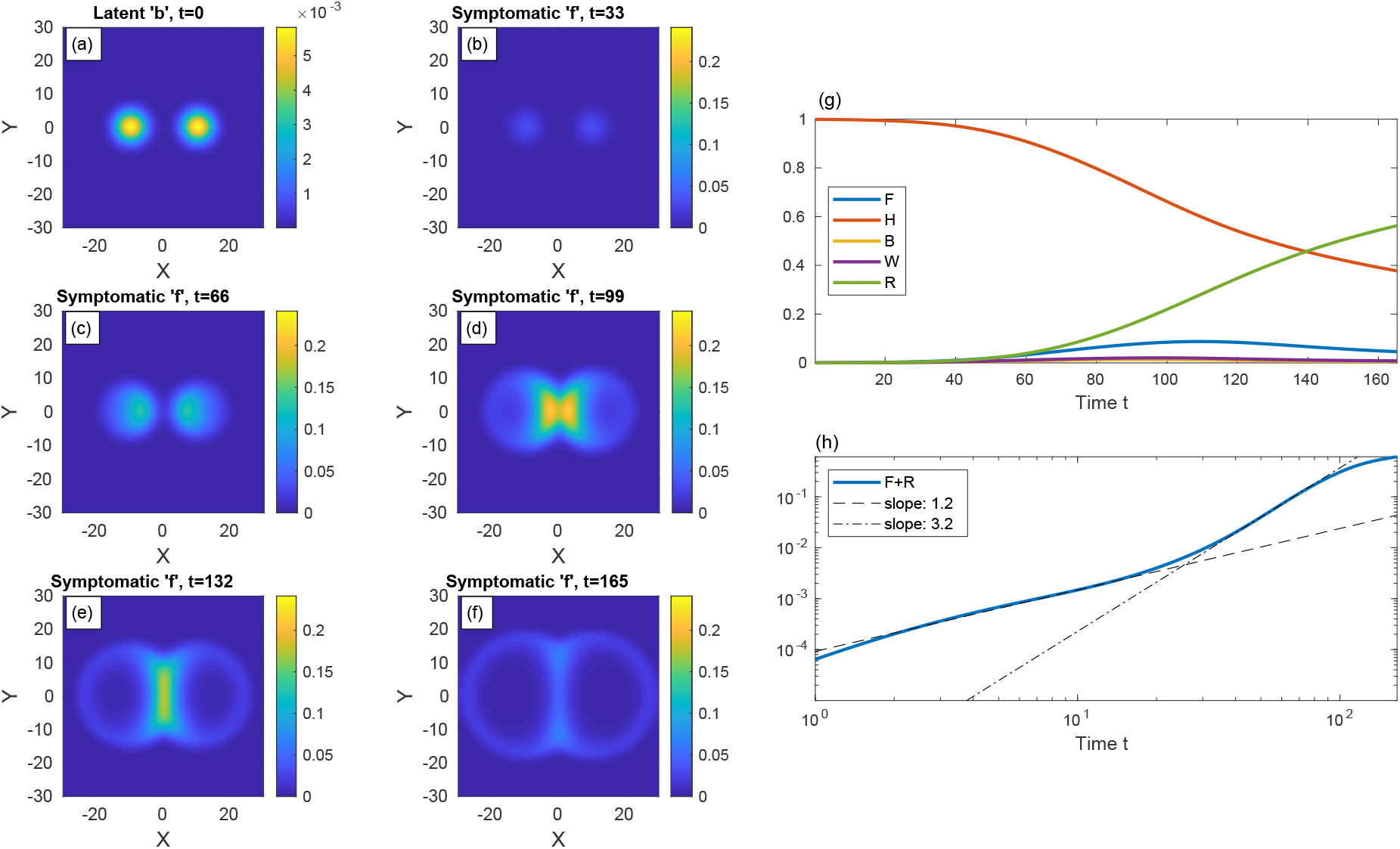
Time evolution of a epidemic starting from two infection centers near a heavily populated region, see (a) (top-left panel). *n* is nonuniform and given by 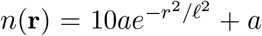, with *ℓ* = 10 and *a* taken such that the spatial average of *n* is 0.2. The global value of *b* is the same as in previous figures, *B* = 10^−3^, and all other populations are initially zero: *w* = *f* = *r* = 0. Panels (b)-(f) show the spread of the symptomatic population ‘*f* ‘as time progresses. The symptomatic population quickly diffuses into the denser region in the center and its density there increases dramatically. The global populations and the sum *F* + *R* are shown in (g) and (h), respectively.

Consider first an infection initiating from around the city center (case (i)), and so we take, as the initial conditions for *b*, several small centers randomly distributed within the city core. Initially, the epidemic consumes non-negligible portion the susceptible (*h*) population within the city core, associated with a substantial growth of *f*, see Figs. 6 (b) and (c). After this initial evolution (*t* ≳ 90) the infection slowly spreads outward by formation of ring-like patterns. Conversely, when the infection initiates from the city outskirts (case (ii), Fig. 7), the pattern is more heterogeneous, and local infection centers grow effectively independently of each other. However, as the epidemic reaches the core of the city, the relatively high density of susceptible population (*h*) allows the *f* -population to keep rising. Hence *F* (*t*), seen in Fig. 7 (g), grows for a longer time and reaches higher values, as compared to case (i) (Fig. 6). The curve *F* (*t*) it is also broader, i.e., we find again the curve flattening effect occurring naturally.

To further investigate this phenomenon, we consider in Fig. 8 (case (iii)) an epidemic that initiates from two larger infection centers on two sides of the city core. We observe that epidemic peak timing and duration is intermediate between cases (i) and (ii). Note the remarkable strong influx of the epidemic towards the densely populated area, as evident from panels (d) and (e); the two growing infection centers merge into one large spot with significantly more symptomatic people. The “late” power-law exponent in this case is ≃ 3.2.

So far we looked at the unperturbed spread of an epidemic. Yet, authorities often use numerous active tools to slow down and/or confine the spread of the disease. Usually people are instructed to stay home for a considerable period, the so-called “quarantine” or “lockdown”. In addition, roads connecting “hotspots” to uninfected regions are sometimes blocked. Under quarantine, there are fewer and less frequent encounters between people implying locally reduced values of *D*_*k*_ and *k*.

To mimic harsh movement restrictions between a city and its surroundings, we impose significantly reduced values of *D*_*k*_ and *k* within a “belt” surrounding the city, which we term “belt quarantine”. In Figs. 9 and 10 we examine the effectiveness of belt quarantine. In both figures, the city is modeled as a region of high population density of radius *ℓ* = 10. In addition, at time *t* = 0, there are small random infection centers surrounding the city. Figure 9 depicts the case of no belt quarantine (uniform *D*_*k*_ and *k*), whereas Fig. 10 represents belt quarantine – between *ℓ* = 10 and *ℓ* = 12 – with values of *D*_*k*_ and *k* reduced to 20% of their background values. As seen from the comparison of the epidemic spreading patterns, with belt quarantine (Fig. 10 (a)-(f)) the infection takes considerably longer time to invade into the densely populated region. The overall effect of the belt quarantine is shown in Fig. 10 (g), where the increase of the global fraction of symptomatic population (*F*) is seen to slow down significantly; e.g., at the longest simulation time we obtain *F* ≃ 9% without quarantine (Fig. 9 (h)), and *F* ≃ 2.6% with belt quarantine (Fig. 10 (h)). The difference in the accumulated infected population, *F* + *R*, is less significant – at the longest simulation time *F* + *R* ≃ 54% without quarantine, and *F* + *R* ≃ 23%.

**FIG. 9.**
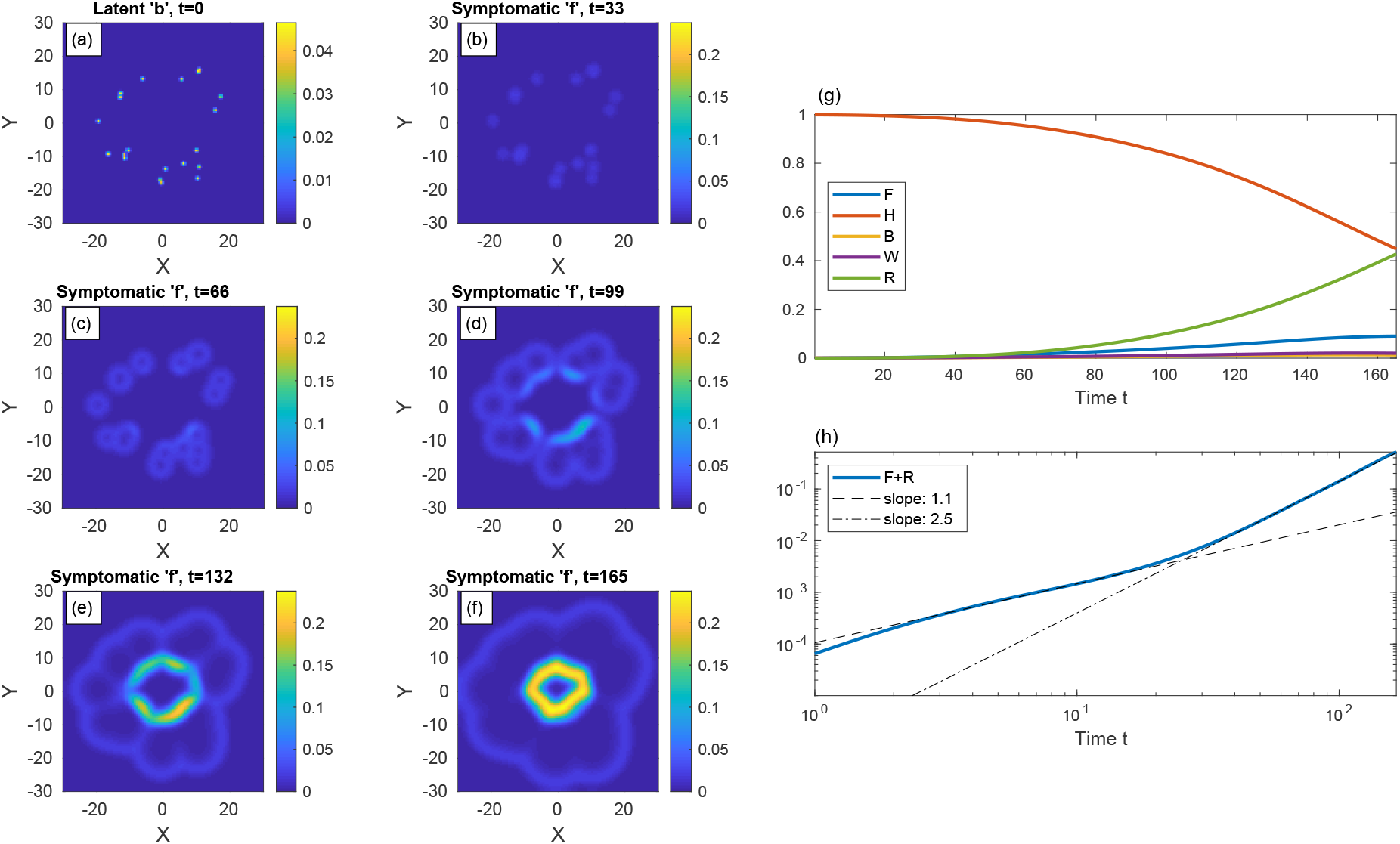
Time evolution of a epidemic starting from multiple random infection centers near a heavily populated region mimicking a city, see (a) (top-left panel). *n* is nonuniform and is given by *n* = 10*a* inside a circle with radius *ℓ* = 10 and *n* = *a* outside of that circle, with *a* taken such that the spatial average of *n* is 0.2. The integrated value of *b* is the same as in previous figures, *B* = 10^−3^. All other populations are initially zero: *w* = *f* = *r* = 0. Panels (b)-(f) depict how the symptomatic population ‘*f* ‘spreads with time and “invades” the city.

**FIG. 10.**
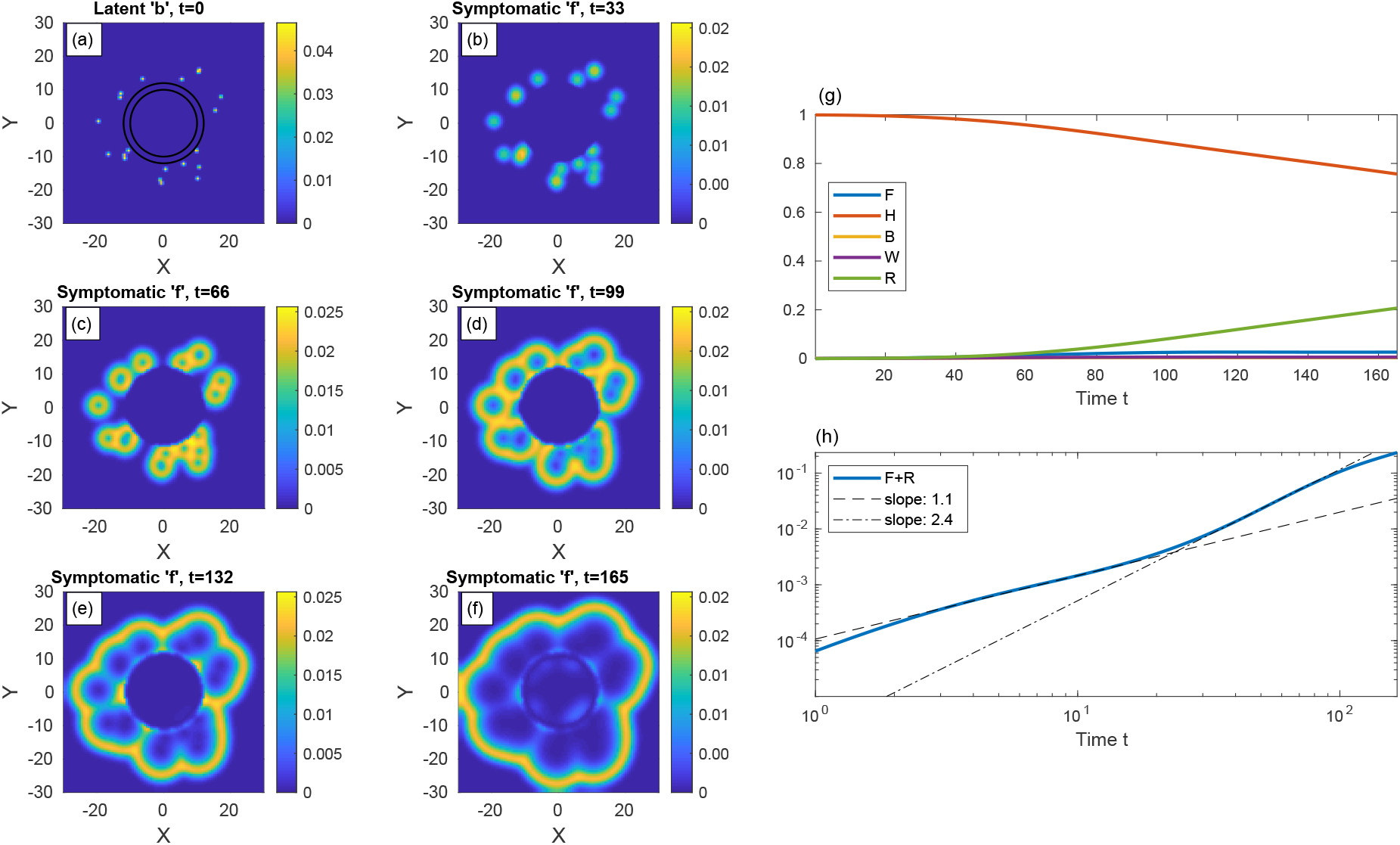
Belt quarantine. Same as Fig. 9 but now the “city” is under protective circumferential “belt” quarantine. Both *D*_*k*_ and *k* are reduced to 20% of their values elsewhere between the two concentric circles (radii *ℓ* = 10 and *ℓ* = 12). The infection spreads quickly within the external area, but penetrates very slowly into the protected region.

Finally, we turn to investigate a neighborhood, which is considered as the epidemic epicenter, within a large urban area. Namely, within the neighborhood there is a high fraction of infected population. The population density *n* is uniform in the whole region of study. In addition to the belt quarantine, we examine here the effect of a more severe measure: lockdown on the whole neighborhood, which we term “area lockdown”. The neighborhood is modeled again as a high population density region of radius *ℓ* = 11. At time *t* = 0, we impose small randomly distributed infection centers within the neighborhood.

Figure 11 depicts the spreading patterns for the case of no quarantine and uniform values of *D*_*k*_ and *k*. Figure 12 represents the effect of belt quarantine between *ℓ* = 10 and *ℓ* = 11. Figure 13 depicts the consequences of area lockdown within the whole neighborhood (circular region of *ℓ* = 11). In both belt quarantine and area lockdown the values of *D*_*k*_ and *k* are reduced to 20% of their background values.

**FIG. 11.**
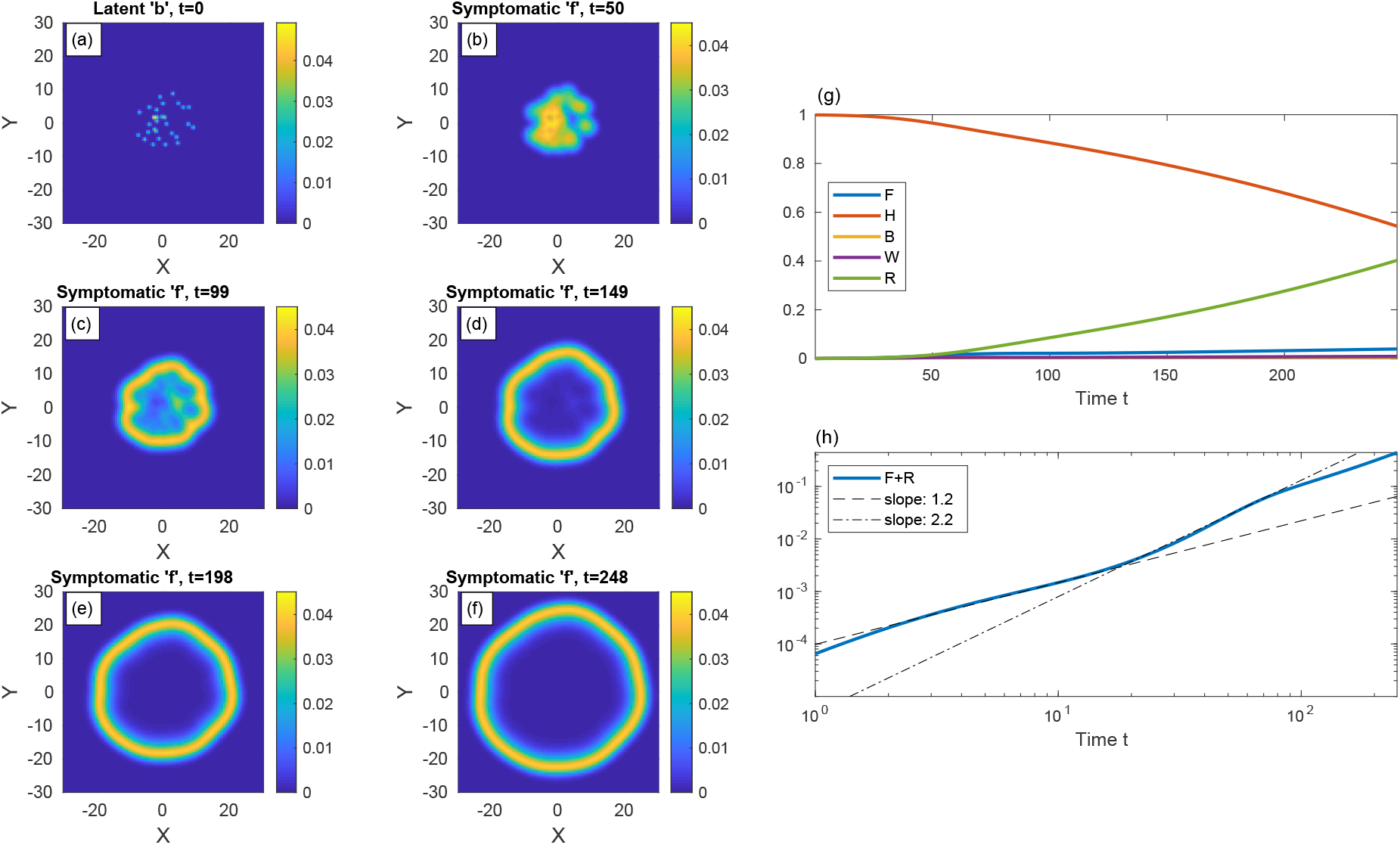
(a)-(f): Time evolution of a epidemic starting from multiple random infection centers with uniform *n*, see (a). The global value of *b* is the same as in previous figures, *B* = 10^−3^. All other populations are initially zero: *w* = *f* = *r* = 0. Both *D*_*k*_ and *k* are uniform. The symptomatic population ‘*f* ‘spreads and forms a ring-like structure that expands in time. (g) and show (respectively) the different global populations and the fraction of population that has been infected until time *t, F* + *R, vs* time.

**FIG. 12.**
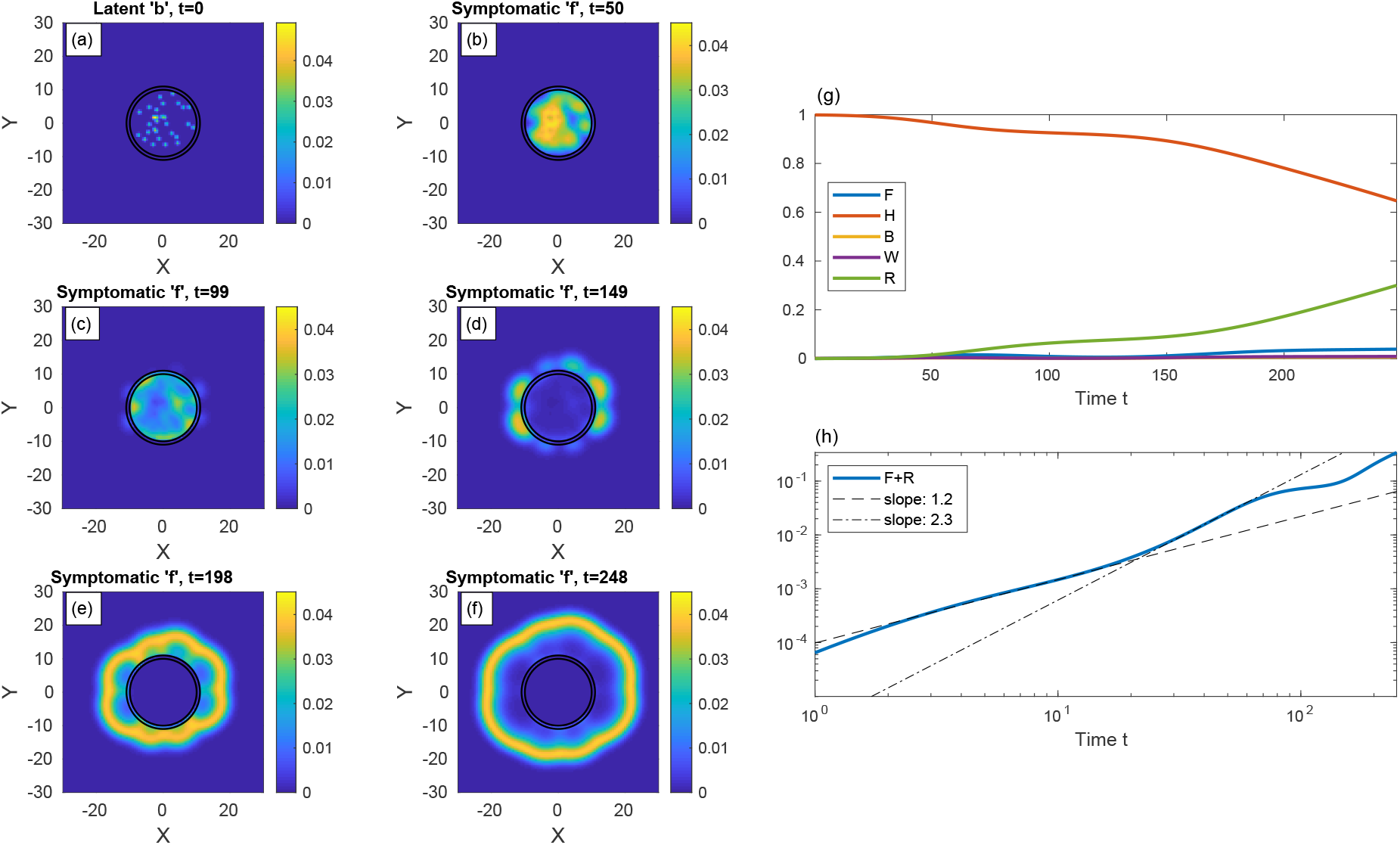
Belt quarantine. Same as in Fig. 11 but now with a protective “belt” quarantine in the region between the two concentric circles (*ℓ* = 10 and *ℓ* = 11). Within the belt, the values of *D*_*k*_ and *k* are 20% of their values in other regions. Initially the infection is confined to the protected region, but at long times it leaks out through the belt and contaminates the exterior. Note the relatively isotropic spreading patterns in the exterior.

**FIG. 13.**
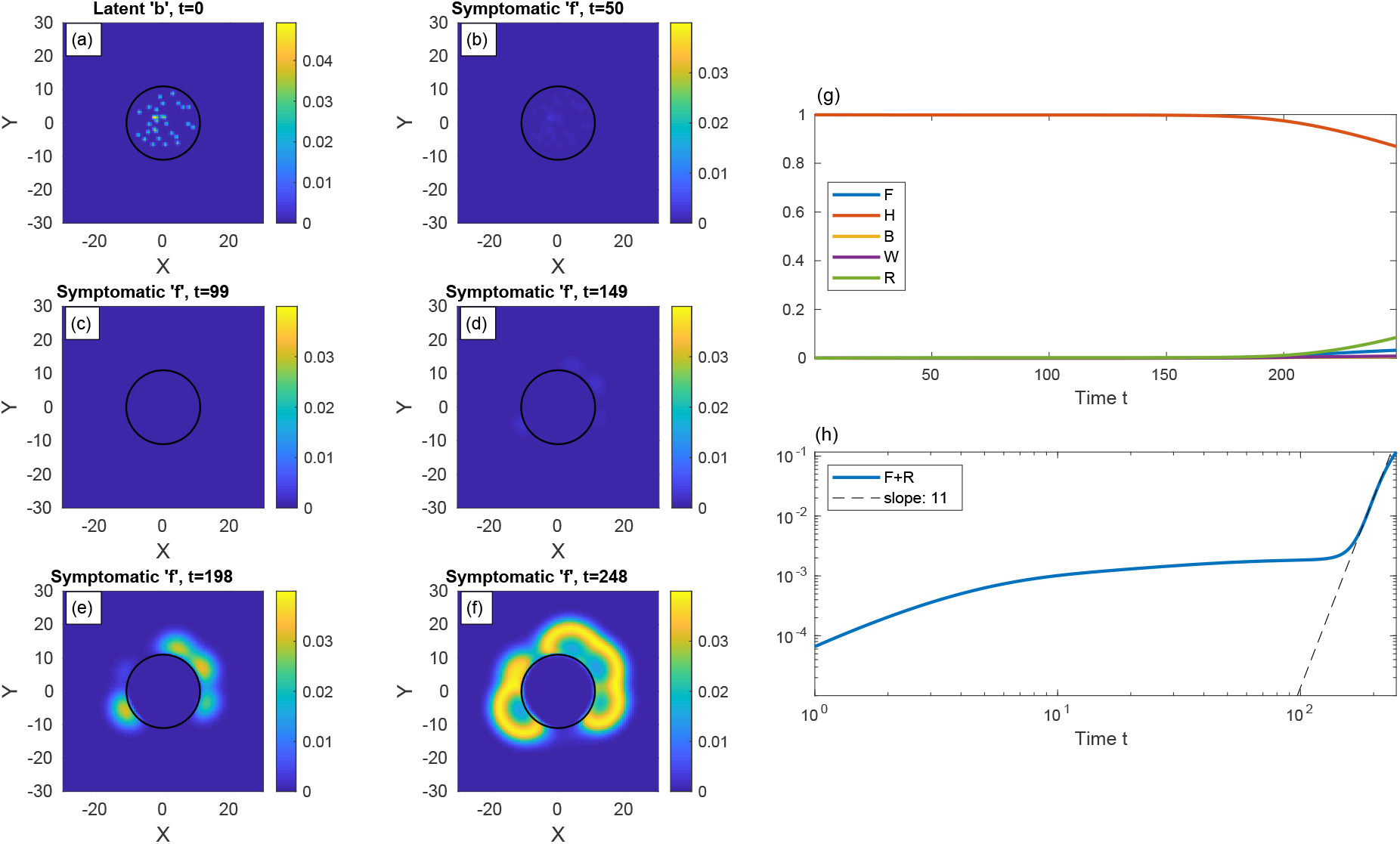
Area lockdown. Same as Fig. 12 but now the quarantine is throughout the whole area of the circle (*ℓ* = 11). Within this quarantined area, the values of *D*_*k*_ and *k* are 20% of their values in other regions. The contamination of the exterior is slower than in Fig. 12. Also note the relatively anisotropic spreading patterns seen at long times in the exterior, as compared to Fig. 12.

The comparison of the epidemic spreading patterns, with belt quarantine (Fig. 12), shows that the escape of the infection from the neighborhood to its surroundings takes longer time in comparison with the no quarantine situation (Fig. 11). However, area lockdown (Fig. 13) is even more efficient than belt quarantine in reducing this escape time. Moreover, it appears that in the case of belt quarantine the late-time spreading pattern is nearly isotropic, while in the case of area lockdown the escape pattern is highly non-isotropic. This occurs since prior to the escape, in the case of belt quarantine the infected population homogenizes rather quickly within the neighborhood, while for area lockdown the slow spreading within the neighborhood prevents this homogenization.

The overall effect of the different quarantine measures on the global populations is shown in panels (g) and (h) of the respective figures. The increase of the global fraction of symptomatic population (*F*) is the smallest under area lockdown; at the longest simulation time, we obtain *F* ≃ 3.9% without quarantine, *F* ≃ 3.8% with belt quarantine, and *F* ≃ 3.2% with area lockdown (relative reduction by 15%). These values are small since, for very slow epidemic evolution, symptomatic people have enough time to recover, such that the “in-flux” of people to this group (from the *w*-population) nearly equals the “out-flux”. A more pronounced effect of the two types of quarantine, which implies on their effectiveness, appears in the accumulated values of infected populations, *F* + *R*: at the longest simulation time we obtain *F* + *R* ≃ 44% without quarantine, *F* + *R* ≃ 33% with belt quarantine, and *F* + *R* ≃ 11.7% with area lockdown (relative reduction by about 70%).

## III. DISCUSSION

An epidemiological spreading model with inherent spatial dependency of the different populations is presented. We take into account nearest-neighbor infection kinetics and show that they lead to diffusion-like terms in the dynamical equations, thereby providing a unified framework for a heterogeneous spread of the epidemic. We show that the complex pattern formation is sensitive to the initial conditions, i.e., to the spatial location of initial infected population, which has important consequences for the total number of infected people. Thus, the observed deviations from the early evolution can be rationalized without assuming time variation in the infection rates, as is customary done in the conventional SEIR-type models.

Our model can naturally describe the flux of infection from a suburban area into a densely populated city, or in the opposite direction. Interestingly, we find that relative “curve flattening” of the infected-symptomatic population (*F*) can naturally occur due to either non-uniform population density, non-uniform distribution of infectious populations, or the combination of both. This may have important implications when comparing the epidemic evolution in different regions or states, where one needs to distinguish between the effects of quarantine measures and population conditions. For example, when comparing Sweden (essentially no quarantine measures) and Israel (severe quarantine measures), conclusions might be hampered.

Importantly, the possibility to mimic in our model spatially-varying and evolving quarantine or lockdown conditions, by using both spatially-dependent (as done in this work) and time-dependent values of *D*_*k*_ and *k*, will allow a quantitative predictive tool for the effectiveness of quarantine measures. This will require complete data sets for the variable population density and the initial “heat maps” for the different types of population defined in our model. Such heat maps are currently produced for COVID-19 by different authorities using extensive testing, and appear occasionally in the media or public websites; some of them resemble our ring patterns [29]. We hope authorities will use this tool in addition to established venues [8, 30] to simulate different lockdown policies for choosing the best exit strategy [31].

## Data Availability

Data may be provided upon request by e-mail.

## Acknowledgments

We are grateful to Ariel Kushmaro for insightful discussions. YT Acknowledges support by the Israel Science Foundation Grant No. 274/19.

